# A cross-sectional audit and survey of Open Science and Data Sharing practices at The Montreal Neurological Institute-Hospital

**DOI:** 10.1101/2022.08.03.22278384

**Authors:** Sanam Ebrahimzadeh, Kelly D. Cobey, Justin Presseau, Mohsen Alayche, Jessie Virginia Willis, David Moher

**Affiliations:** Centre for Journalology, Ottawa Hospital Research Institute, Ottawa, Canada; University of Ottawa Heart Institute, Ottawa, Canada; School of Epidemiology and Public Health, University of Ottawa, Ottawa, Canada; Faculty of Medicine, University of Ottawa, Ottawa, Canada

**Keywords:** Open Science, Data Sharing, Audit, Survey, Facilitators and barriers

## Abstract

**Objectives:** To audit all publications produced by Montreal Neurological Institute-Hospital researchers regarding open science practices and to survey Neuro-based researchers about barriers and facilitators to data sharing.

**Setting, design and participants:** In the first study, we retrieved 313 unique publications and collated all Neuro publications from 2019 and extracted information from each article pertaining to data sharing and other open science practices. We included all empirical papers and pre-prints that were reported in English. In the second study, one hundred twenty-four participants (out of 553) completed the survey, a response rate of 22.42%. We surveyed all Neuro researchers.

**Primary and secondary outcomes:** for the audit we examined data sharing and open science practices. For the survey, we asked participants about their data sharing practices.

**Results:** We found that 66.5% of these publications (n=208) included a data sharing statement. Overall, 74.5% (n=155) of articles had data that was publicly available. When examining broader open science practices, rates of compliance tended to be lower. For example, 94.9% (n=297) of publications failed to register a protocol. Among participants who had published a first or last authored paper in the past year, most participants, 53 of 74 (71.62%), reported that they had openly shared their research data. Less than half of the participants 37.50% (n=45) reported having engaged in training related to data sharing within the last 12 months.

**Conclusion:** We found that half of all publications included in the audit shared data. Participants indicated an appetite for resources for learning about data sharing signaling a willingness to perform better.

**Strengths and limitations of this study:**

- To serve as a baseline to benchmark for improvements in data sharing and other open science practices
- To measure progress over time.
- The results of the study cannot be generalized.
- It is hard to measure changes in the community.

## Introduction

Open science refers to the practices of making research outputs openly available for others to use and build upon. Open science can be seen as an alternative to the typically ‘closed’ model of science which tends to stress concern over intellectual property and controlling access to scientific knowledge and data (1). Benefits to open science include increased transparency, the ability to replicate research, equity in access to research information, reputational gains and increased chances of publication (2). Recognizing the value of open science, many stakeholders have begun to implement policy mandates to see that open science practices are being implemented in the community. This includes the creation of federal roadmaps and policies, organization policies, and funder policies, such as Canada’s Roadmap to Open Science (3), the second French national plan for open science (4), the United Nations Educational, Scientific and Cultural Organization’s open science policy (5), the European Union’s open science policies (6), and others.

Beyond these stakeholders, academic institutions have an important role in implementing open science (1). Despite their seemingly obvious role, academic institutions have tended not to feature in discussions around implementing open science. Indeed, academic institutions have been previously criticized for not playing more of a role in addressing issues of research reproducibility (7) which is conceptually related to open science. The reality is that few academic institutions have strong transparent processes in place to encourage open science. Despite this, research institutions are uniquely positioned to help contribute to defining incentives in research and to valuing open science practices. Shifting researchers’ behavior towards openness will require education, training, as well as a culture shift. Research institutions could provide the environment of this training to verify common understanding of open science and to help shift culture. Inaction by research institutions to play their role in implementing open science will have downstream consequences for how research is disseminated.

In Canada, a notable exception is the Montreal Neurological Institute-Hospital (henceforth called the ‘Neuro’) which has committed to becoming an open science institute. Having made the scientific and cultural decision to become ‘open’ after a significant consultation and buy-in process (8), the Neuro is now positioned to implement open research practices. The Neuro has made a structural decision to run a focused implementation program wherein a small set of open science practices are initially focused on, and additional practices then added in an incremental manner.

At present there is a focus on data sharing, namely making the data underlying the results reported in a given publication publicly available for others to use and build upon, or to verify the work reported. The rationale of choosing data sharing from the potential open research practices was: 1. few researchers have the skills required to share their data and most researchers are not trained in data sharing(9); 2. Data sharing has the potential to lead to novel discovery and enhances the transparency of disseminated research findings; 3. the largest government health funder in Canada, Canadian Institutes of Health Research, recently announced new polices on data management (10). By adopting data sharing now, the Neuro stands to be ahead of incoming mandates and may offer insight for others to follow. Finally, while some data sharing infrastructure exists, a significant investment is being made at the federal level to build capacity for Canadian researchers (11) and (12)

This report describes the results of two studies examining data sharing practices at the Neuro. In the first study, we audited all publications produced by Neuro researchers in 2019. In the second study, we surveyed Neuro-based researchers about barriers and facilitators to data sharing. The results will provide the Neuro with a better understanding of barriers and facilitators to data management and sharing and will identify educational needs related to data sharing that can be implemented to address barriers. Further, the audit of publications can be used to benchmark for improvements over time and to monitor change.

## Method

### Transparency statement

We used the STROBE guideline to inform our reporting of the audit (13) and the CHERRIES guideline to inform reporting of the survey (14)

### Study 1: A cross-sectional audit of Neuro publications

#### Search strategy

We identified the Neuro publications produced in 2019 by searching the Web of Science (WoS) (15) and using its meta-data to capture the Neuro’s output including searching two preprint servers (MedRxiv (16) and bioRxiv (17). The search strategy was developed by trained information specialists and librarians (Sanam Ebrahimzadeh and Alex Amar-Zifkin). For the full search strategy please see appendix 1.

#### Patient and Public Involvement

Patients or the public were not involved in the design, or conduct, or reporting, or dissemination plans of our research.

#### Eligibility Criteria

We aimed to include all research papers disseminated by researchers based at the Neuro in 2019. We included research papers within any field. We used the last listed date of publication on each paper to determine if the publication fit within this timeframe. We included all research publications irrespective of the role of the Neuro based author (e.g., including trainees, graduate and postdoctoral students, early career researchers and more established researchers). We included all publications where a Neuro-based author was listed, irrespective of where they were named on the author byline. We included publications in English only. We included publications in traditional peer-reviewed journals as well as those on preprint servers.

#### Screening and extraction

DistillerSr (18) was used to manage study records retrieved by the search. We obtained all full-text documents, and two reviewers independently screened these records against our inclusion criteria for included articles, we then extracted basic epidemiological information from each included study, including the names of the Neuro based author(s), the outlet of publication, and we classified articles based on their study design and content area. In addition, we extracted information related to data sharing practices. This included information on whether the publication contained a data sharing statement, whether or not data sharing occurred, and if so, what format and tools were used for this sharing. In addition, we extracted information on a range of other open science practices as per table 1.

**Table 1.** Other open science practices examined in the audit

| **Open Science Practice** | **Description** |
| --- | --- |
| **Open access** | Articles were published in a format that made them free to access and build upon |
| **Study registration** | Articles report having created a time-stamped read only version of the study protocol which was shared prior to data collection |
| **Preprints** | Articles were a preprint, or referenced that the publication had a linked preprint |
| **Reporting Guideline** | Articles reported adhering to a reporting guideline checklist |
| **Reference to a protocol** | Articles linked to a study protocol (but this protocol was not formally registered) |
| **Ethics approval** | Articles indicated ethics approval was obtained/waived (or not required) |
| **Conflict of interest** | Articles included a statement of conflicts of interest |

#### Data analysis

Once all extracted data were in agreement (i.e. discrepancies between assessors resolved) the complete dataset for all included articles was exported from DistillerSR into SPSS 28 (19) where data were cleaned. We presented the total number of included articles, and basic descriptive analyses for all items extracted using count data and percentages. Also, we used Unpaywall (20) to check the open access status of the articles captured in our sample. To do so, we inserted the DOIs we extracted from the included articles into the Unpaywall tool

### Study 2: A cross-sectional online survey of Neuro-based researchers Survey

#### Sampling

The survey was closed (i.e., only open to those we invited) and was administered using SurveyMonkey software (21).

#### Survey Items

After providing informed consent, participants were presented with a 14-item survey (See Appendix 2). The survey was custom-built but draws on items previously reported by Van Panhuis (22). We presented participants with a series of questions regarding their willingness to share data that were developed using the 14 domains of the Theoretical Domains Framework (TDF) to help structure the survey items pertaining to barriers and facilitators to data management and actual data sharing. The TDF was created to better understand health professional behavior and is an integrated theoretical framework (23). Participants were asked to indicate their level of agreement with each statement on a Likert scale of 1 (strongly disagree) to 5 (strongly agree) was used to allow the participants to express how much they agree or disagree with each item. The Neuro ‘Open Science Grassroots committee’, a group of Neuro-based researchers targeting improvements in open science at the institution and beyond (24), reviewed and provided feedback on the survey which we incorporated. The survey was also piloted by three researchers prior to dissemination for clarity.

#### Survey recruitment

We included all the Neuro’s currently employed graduate students, postdoctoral candidates, research support staff, and independent investigators. The list of researchers was provided to us by the Neuro’s human resources. Participants were invited to complete the survey by Dr. Guy Rouleau, Director of the Neuro, via email, using a standard recruitment script. Participants were asked to complete informed consent online. The consent form described the aims of the study, specify that data collected would be anonymous, and detail our data management plan which includes making data openly available. Completion of the survey was by as implied consent.

Participants were originally sent the survey on Sept 20^th^, 2021, with a standardized reminder sent after one week (September 27^th^, 2021). Some targeted internal email strategies were also implemented to help maximize the response rate. We closed the survey on November 3^rd^, 2021.

#### Data analysis

Data was analyzed using IBM SPSS 28. We also report the completion rate for individual items, and report frequencies and percentages, or means and standard deviation, for each of the survey items. We report comparative data, using a Chi square test, illustrating findings by researcher category and gender.

For open-ended survey questions, we conducted a thematic analysis (25) of responses provided. Here, two researchers familiarized themselves with the responses to the qualitative items (questions 8, 13, and 14, see appendix 1). Then, they each, independently, coded responses. Codes were discussed and iteratively reviewed until there was consensus. Then, key themes were identified, and codes grouped within these. Themes were defined and described in the results section, below.

## Results

### Study 1: A cross-sectional audit of Neuro publications

#### Sample

We initially identified 623 publications from our search. We then removed 15 duplicates. We screened a total of 608 unique references, 313 publications met our inclusion criteria (See Figure 1). Of the retrieved 623 publications, about half were not research outputs. Epidemiological characteristic of included publications is provided in Table 2.

**Table 2.**
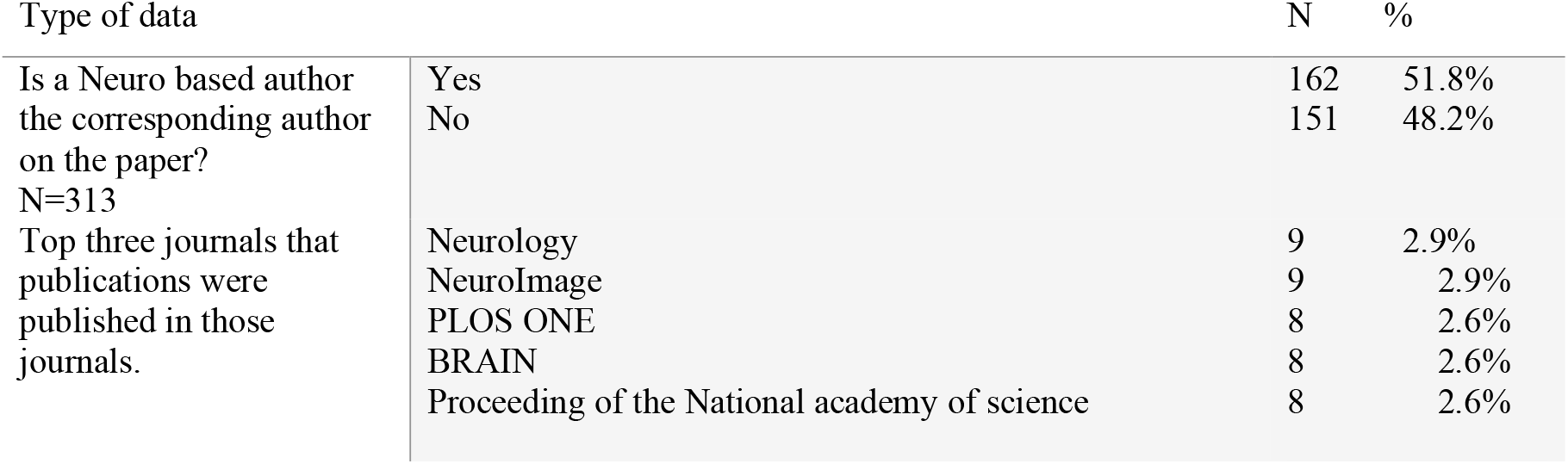
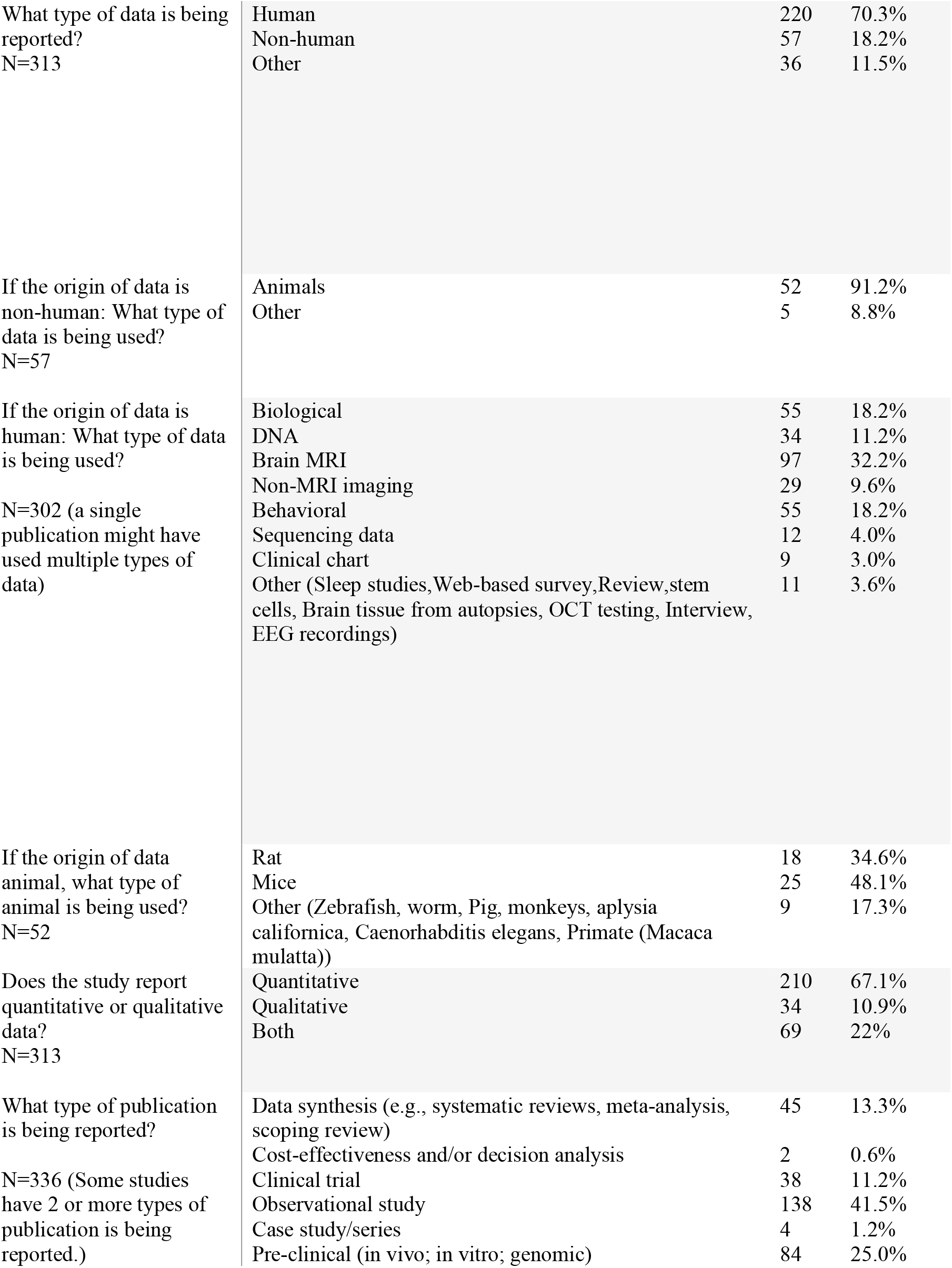
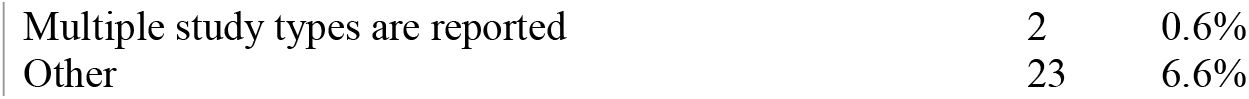
Epidemiological characteristics of include articles

| Type of data |  | N | % |
| --- | --- | --- | --- |
| Is a Neuro based author the corresponding author on the paper?<br>N=313 | Yes | 162 | 51.8% |
|  | No | 151 | 48.2% |
| Top three journals that publications were published in those journals. | Neurology | 9 | 2.9% |
|  | NeuroImage | 9 | 2.9% |
|  | PLOS ONE | 8 | 2.6% |
|  | BRAIN | 8 | 2.6% |
|  | Proceeding of the National academy of science | 8 | 2.6% |
| What type of data is being reported?<br>N=313 | Human | 220 | 70.3% |
|  | Non-human | 57 | 18.2% |
|  | Other | 36 | 11.5% |
| If the origin of data is non-human: What type of data is being used?<br>N=57 | Animals | 52 | 91.2% |
|  | Other | 5 | 8.8% |
| If the origin of data is human: What type of data is being used?<br>N=302 (a single publication might have used multiple types of data) | Biological | 55 | 18.2% |
|  | DNA | 34 | 11.2% |
|  | Brain MRI | 97 | 32.2% |
|  | Non-MRI imaging | 29 | 9.6% |
|  | Behavioral | 55 | 18.2% |
|  | Sequencing data | 12 | 4.0% |
|  | Clinical chart | 9 | 3.0% |
|  | Other (Sleep studies, Web-based survey, Review, stem cells, Brain tissue from autopsies, OCT testing, Interview, EEG recordings) | 11 | 3.6% |
| If the origin of data animal, what type of animal is being used?<br>N=52 | Rat | 18 | 34.6% |
|  | Mice | 25 | 48.1% |
|  | Other (Zebrafish, worm, Pig, monkeys, aplysia californica, Caenorhabditis elegans, Primate (Macaca mulatta)) | 9 | 17.3% |
| Does the study report quantitative or qualitative data?<br>N=313 | Quantitative | 210 | 67.1% |
|  | Qualitative | 34 | 10.9% |
|  | Both | 69 | 22% |
| What type of publication is being reported?<br>N=336 (Some studies have 2 or more types of publication is being reported.) | Data synthesis (e.g., systematic reviews, meta-analysis, scoping review) | 45 | 13.3% |
|  | Cost-effectiveness and/or decision analysis | 2 | 0.6% |
|  | Clinical trial | 38 | 11.2% |
|  | Observational study | 138 | 41.5% |
|  | Case study/series | 4 | 1.2% |
|  | Pre-clinical (in vivo; in vitro; genomic) | 84 | 25.0% |

|  |  |  |
| --- | --- | --- |
| Multiple study types are reported | 2 | 0.6% |
| Other | 23 | 6.6% |

**Figure 1.**
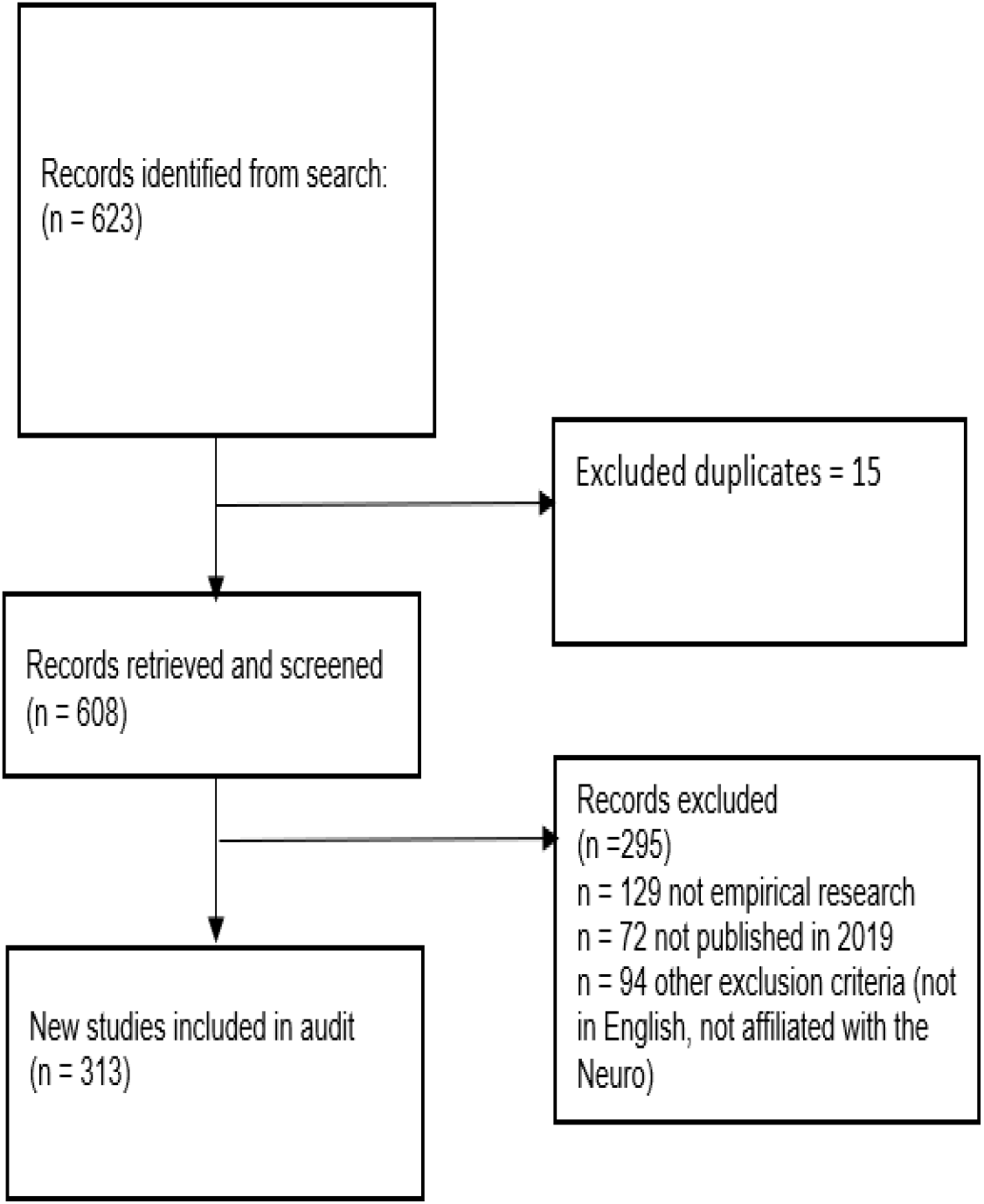
Study flow of records and studies remaining and excluded publications.

In about half of the included publications (52%; n = 162), the Neuro based author was the corresponding author on the paper. Neurology 2.9% (n=9), NeuroImage 2.9% (n=9), PLOS One 2.6% (n=8), Brain 2.6% (n=8) and Proceeding of the National academy of science 2.6% (n=8) were the most prevalent journals that Neuro authors published in (see Table 2). The number of authors in the included papers ranged between 1-374 authors. Studies with more than 100 authors were typically a systematic analysis, such as GBD 2016 Traumatic Brain Injury and Spinal Cord Injury Collaborators. Global, regional, and national burden of traumatic brain injury and spinal cord injury, 1990-2016: a systematic analysis for the Global Burden of Disease Study 2016 (26). The median number of authors was eight. In addition, we investigated the number of authors with a Neuro affiliation. We found that 31% (n=104) of the publications had a Neuro affiliated author. The number of authors with the Neuro affiliation ranged between 1-12 in the included publications. The median number of the Neuro affiliated authors on a given publication was two.

Seventy percent of the included papers reported on clinical research (n=220), while 18.2% (n=57) reported non-human data, and 11.5% (n=36) used another form of data, such as literature review, protocol, both clinical research and non-human and simulation analyses.

When we examined the type of clinical data shared, we found that biological data (e.g., implementation of an antibody) was used in 18.2% (n=55) of publications, DNA in 11.2% (n=34), brain MRI in 32.2% (n=97), non-MRI imaging in 9.6% (n=29), behavioral data in 18.2% (n=55), sequencing data in 4.0% (n=12) and clinical chart in 3% (n=9) of publication. Sleep studies, web-based surveys, stem cells, brain tissue from autopsies, OCT testing, interviews, EEG recordings were used in the “other” 3.6% (n=11) publications.

We also examined data sharing of non-human research publication. We found that animal data was being used in 91.2% (n=52) publications, five (8.8%) publications used other data (e.g., radiology technology, model). Mice 48.1% (n=25) and rats 34.6% (n=18) were being used in most of the publications. Sixty-seven percent (n=210) of the publications reported quantitative data and 10.9% (n=34) reported qualitative data. Twenty-two percent (n=69) of publications reported both quantitative and qualitative data. Forty percent of publication (n=138) were observational studies.

For full details of types of data, please see table 2.

#### Data sharing audit findings

Complete results of our data sharing audit are reported in Table 3; here we report key findings. More than half of the publications (66.5%; n=208) included a data sharing statement. Two-thirds of the data sharing statements were found at the end of the paper in a ‘declaration’ section of the publication (66.0%; n=139/208). Other statements were found in the manuscript methods section 19.7% (n=41), in the manuscript results section 2.9% (n=6), or in other sections 26.0% (n=54) such as the front page, abstract, Dryad, or the Supplementary Materials section of the publication. Of the papers that had a data sharing statement, we were able to download 74.5% (n=155/208) of data from the included publications directly. Of the entire sample of articles included, 49.5%; 155/313) of the papers had openly available data.

**Table 3.** Data sharing evaluation

| Data sharing evaluation |  | N | % |
| --- | --- | --- | --- |
| Did the paper include a data sharing statements?<br>N=313 | Yes | 208 | 66.5% |
|  | No | 104 | 33.2% |
|  | Unclear (broken link) | 1 | 0.3% |
| Where did you find the data sharing statement?<br>N=208(the number of manuscripts with data sharing) | In the manuscript Methods section. | 41 | 19.7% |
|  | In the manuscript Results section. | 6 | 2.9% |
|  | In a declaration section. | 139 | 66.9% |
|  | Other (e.g. Front page, Abstract, By request, Dryad links throughout Supplementary Material section) | 54 | 26.0% |
| Can you directly access, download, and open the data? (from articles that had data sharing statements, n=208) | Yes | 155 | 74.5% |
|  | No | 46 | 22.1% |
|  | Other (e.g., By registration, Error page) | 7 | 3.4% |
| If No, does the statement specify to contact the author to gain access to the data? (N=46) | Yes | 27 | 58.7% |
|  | No | 14 | 30.4% |
|  | Other (Registration required, must request an account, request through vivli platform, says attached at bottom of article) | 5 | 10.9% |
| Where does the publication indicate the shared data is located?<br>N=182 | Journal website | 120 | 66.0% |
|  | Other (NeuroVault, Zenodo, github, Genbank, NITRC/ABIDE, bioRxiv website, Dryad) | 30 | 16.5% |
|  | Institutional repository | 17 | 9.3% |
|  | Open Science Framework | 8 | 4.4% |
|  | Figshare | 7 | 3.8% |
| In what format is data shared?<br>N=204 | PDF | 61 | 30.0% |
|  | Word | 59 | 29.0% |
|  | Excel | 29 | 14.2% |
|  | HTML | 2 | 0.9% |
|  | R | 4 | 1.9% |
|  | SPSS | 1 | 0.5% |
|  | Other (3D view, Python Scripts, CSV TIFF, tar.gz, multiple histological datasets, zip, wav, Matlab, avi (video), txt, js, HTTP, mp4, png, table, ATAV) | 48 | 23.5% |
| Does the data published have a unique digital identifier?<br>N=155 | Yes | 45 | 29.0% |
|  | No | 103 | 66.5% |
|  | Unclear (e.g., Can't open the link) | 4 | 2.6% |
|  | Other (the same DOI as the paper itself) | 3 | 1.9% |

When examining publications with data sharing statements that did not have openly available data, 58.7% (n=27 of 46) of them specified to contact the author to gain access to the data. Eleven percent (n=5) of the publications identified other ways to access the data (e.g., registration to a site required, request through the Vivli platform which gives access to anonymized individual participant-level data (IPD) or the cleaned raw data that is collected during a clinical trial (27). Thirty percent (n=14) of these publications did not specify a way to gain access to the data.

When data was shared openly it was done so on: journal websites (n=120, 66.0%), the Open Science Framework (n=8, 4.4%), Institutional repository (n=17, 9.3%), Figshare (n=7, 3.8%), or using other platforms (n=30, 16.5%). Data was shared mostly in PDF format (n=61, 30.0%), Word format (n=59, 29.0%), and Excel format (n=29, 14.2%). Most of the data 66.0% (n=103) shared was published without a unique identifier, with just 29.0% publications (n=45) providing a Digital Object Identifier. It was unclear for 2.6% (n=4) publications if a DOI was used.

#### Open scripts and materials

We found that most publications 75.7% (n=237) indicated that there is no analysis script and/or statistical analysis plan (SAP) available. Twenty percent (n=64) of the publications had analysis/SAPs available. Four (1.3%) publications stated that the analysis scripts and statistics data are available upon request. Two percent (n=8) were other (e.g. available from the corresponding author on reasonable request). The analysis scripts were available via a personal or institutional webpages (n=9, 11.5%), a journal webpage (n=16, 20.5%), supplementary information hosted by the journal or a pre-print server (n=16, 20.5%), an online third party repository (e.g., OSF, Figshare, etc.) (n=26, 33.4%).

When publications shared an analysis script, the vast majority, 93.7% (n=60/64), could be downloaded directly. There was only one publications (n = 64; 1.6%) where we could not download the analysis script.

We defined “Materials availability” as sharing materials used that were to conduct the study. (e.g., such as video of Cognitive Behavior Therapy for psychological interventions; surveys; cell lines; reporting checklist(s), supplementary files, Gene banks). We found that (n=137, 43.8%) of the publications indicated that study materials were available. Less than one percent (n=2) of the publications indicated that the materials were available upon request. Materials were made available via the journal publication webpage (n=51, 30.9%), supplementary information hosted by the journal (n=51, 30.9%), using personal or institutional webpages (n=15, 9.2%), or via an online third-party repository (e.g., OSF, Figshare) fourteen percent (n=23). A small number of articles indicated materials were available by request (n=6, 4.4%). Six presents of materials (n=9) could not be downloaded directly, 92% (n=126) of materials in the publications could be downloaded, and two (1.5%) publications did not use in their publications. Both publication were review (28) and (29)

All of these results are included in Table 4.

**Table 4.**
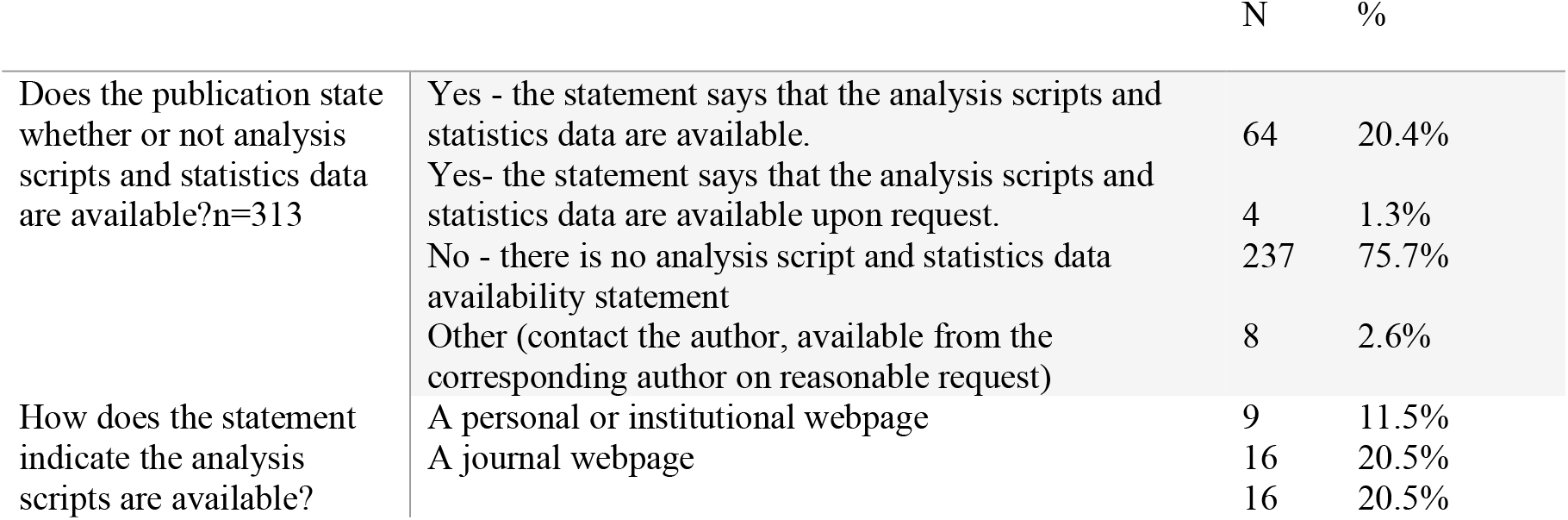
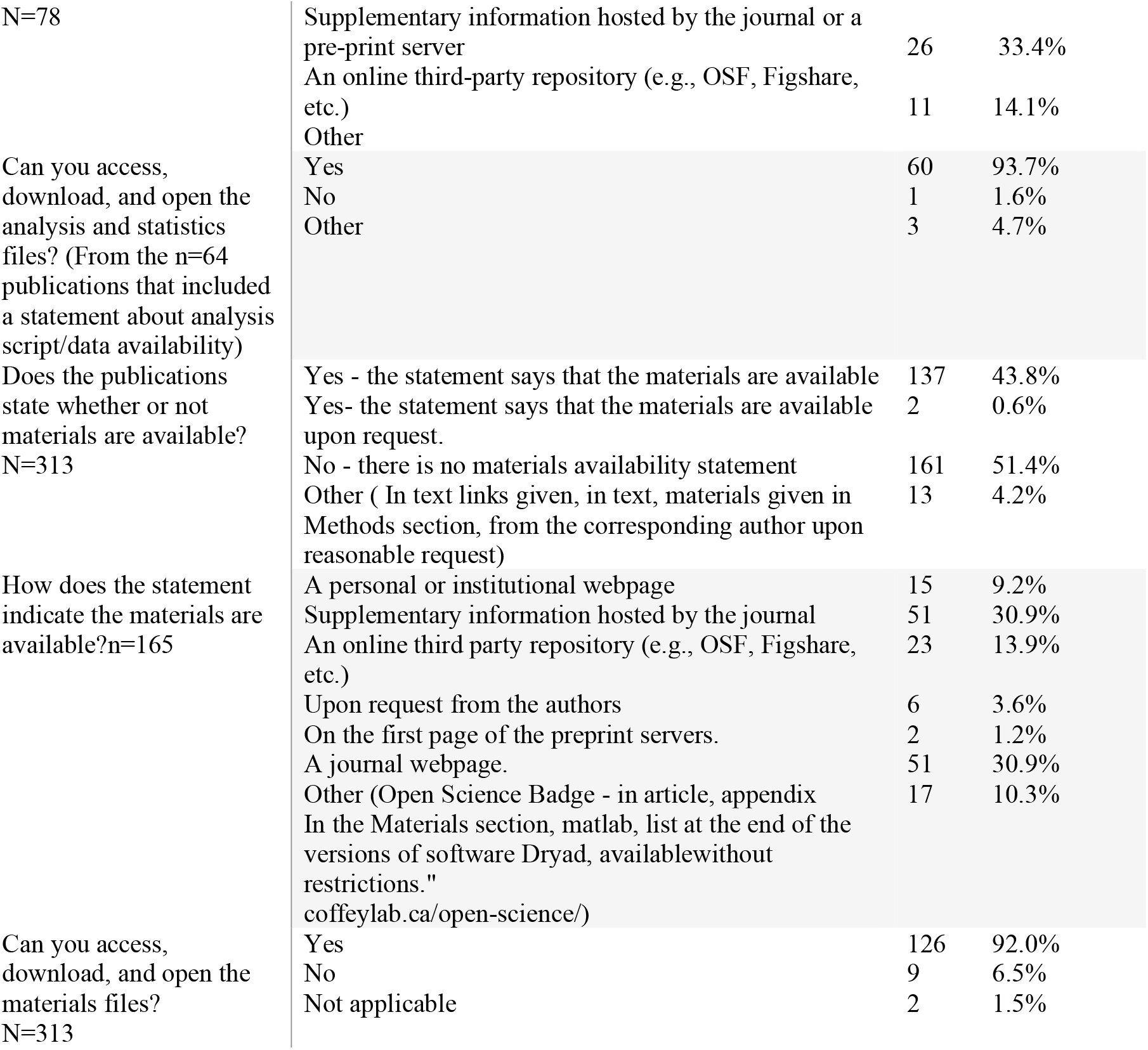
Analysis scripts and material availability

#### Transparency and open science practices

Results from our audit of transparency and open science practices are reported in Table 5. Fifteen percent (n=47) of the publications did not mention a conflict-of-interest statement, 15% (n=47) indicated that there were one or more conflicts of interests and 68.8% (n=215) indicated that there was no conflict of interest. In addition, we investigated whether the publications reported funding and, if so, funding sources. Most of the publications 88% (n=296) reported the funding statements. The primary sources of funding were the federal public funder, the Canadian Institutes of Health Research (CIHR; n=149) and the provincial funder, the Fonds de Recherche du Québec – Santé (FRQS; n=71).

**Table 5.**
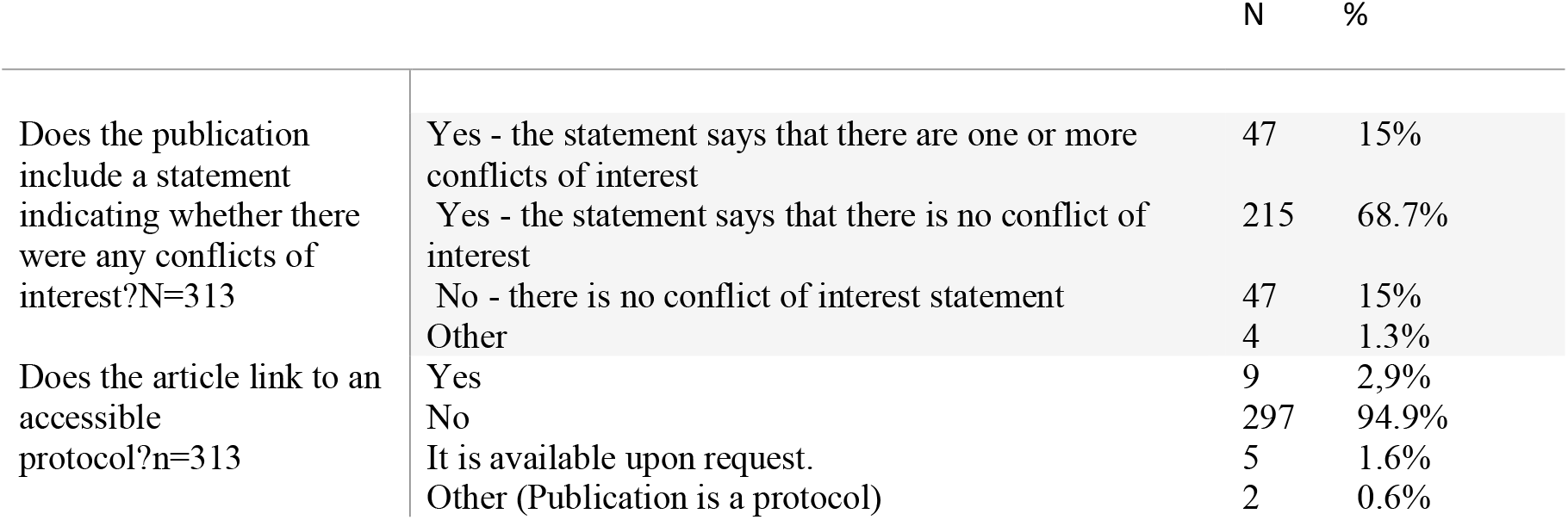
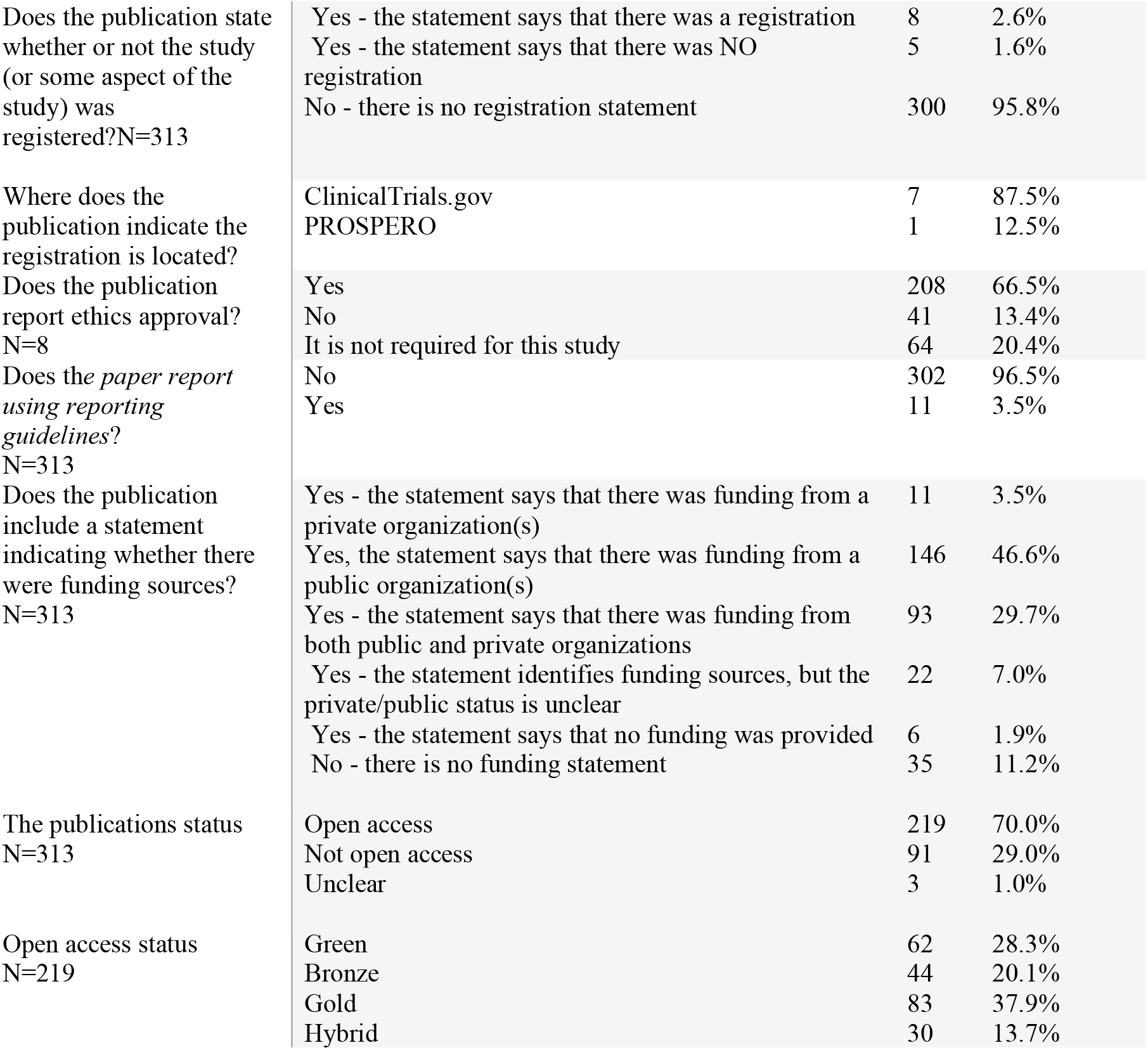
Open access evaluation

|  |  | N | % |
| --- | --- | --- | --- |
| Does the publication include a statement indicating whether there were any conflicts of interest?N=313 | Yes - the statement says that there are one or more conflicts of interest | 47 | 15% |
|  | Yes - the statement says that there is no conflict of interest | 215 | 68.7% |
|  | No - there is no conflict of interest statement | 47 | 15% |
|  | Other | 4 | 1.3% |
| Does the article link to an accessible protocol?n=313 | Yes | 9 | 2,9% |
|  | No | 297 | 94.9% |
|  | It is available upon request. | 5 | 1.6% |
|  | Other (Publication is a protocol) | 2 | 0.6% |
| Does the publication state whether or not the study (or some aspect of the study) was registered?N=313 | Yes - the statement says that there was a registration | 8 | 2.6% |
|  | Yes - the statement says that there was NO registration | 5 | 1.6% |
|  | No - there is no registration statement | 300 | 95.8% |
| Where does the publication indicate the registration is located? | ClinicalTrials.gov | 7 | 87.5% |
|  | PROSPERO | 1 | 12.5% |
| Does the publication report ethics approval? N=8 | Yes | 208 | 66.5% |
|  | No | 41 | 13.4% |
|  | It is not required for this study | 64 | 20.4% |
| Does the *paper report using reporting guidelines?* N=313 | No | 302 | 96.5% |
|  | Yes | 11 | 3.5% |
| Does the publication include a statement indicating whether there were funding sources? N=313 | Yes - the statement says that there was funding from a private organization(s) | 11 | 3.5% |
|  | Yes, the statement says that there was funding from a public organization(s) | 146 | 46.6% |
|  | Yes - the statement says that there was funding from both public and private organizations | 93 | 29.7% |
|  | Yes - the statement identifies funding sources, but the private/public status is unclear | 22 | 7.0% |
|  | Yes - the statement says that no funding was provided | 6 | 1.9% |
|  | No - there is no funding statement | 35 | 11.2% |
| The publications status N=313 | Open access | 219 | 70.0% |
|  | Not open access | 91 | 29.0% |
|  | Unclear | 3 | 1.0% |
| Open access status N=219 | Green | 62 | 28.3% |
|  | Bronze | 44 | 20.1% |
|  | Gold | 83 | 37.9% |
|  | Hybrid | 30 | 13.7% |

Most publications 94.9% (n=297) did not link to an accessible protocol; only 2.9% (n=9) did. 1.6% (n=5) of the publications reported that a protocol was available upon request, and two (0.6%) publications were actual reports of publication protocols.

Most publications 95.8% (n=300) did not include a study registration statement. Only 2.6% (n=8) of the publications indicated registration, 1.6% (n=5) of them indicated that there was no registration. Most registrations 87.5% (n=7) were for clinical trials referring to registration on ClinicalTrials.gov; 12.5% were located on the International Prospective Registry of Systematic Reviews, the PROSPERO database (n=1).

For studies in which ethics approval was considered approval, 66.6% (n=208) reported this information. Ethics approval was not relevant or not required for 20.4% (n=64) publications. Most publications 96.5% (n=302) did not report using a reporting guideline to improve the completeness and transparency of the completed research, only 3.5% (n=11) publications explicitly mentioned adherence to reporting guidelines

We found that 70.0% (n=219) of the articles were open access, while the 29.0% (n=91) were not openly available. Data were missing for 1.0% (n=3) of the journals. Among the studies that were openly available, 37.9% (n=83 of 219) were in ‘gold’’ (i.e., the final published version of your article (or Version of Record) is immediately permanently and freely available online) open access journals. Thirteen percent (n=30 of 219) were published in hybrid journals, 20.1% (n=44 of 219) were in ‘bronze’ (i.e., the final published version of your article is free to read on the publisher page but without a creative commons license) journals, and 28.3% (n=62 of 219) were made available green (i.e., when you place a version of the your manuscript into a repository, often after an embargo, making it freely accessible for everyone)open access via a repository. See table 5.

### Study 2 A cross-sectional online survey of Neuro-based researchers Survey

#### Participants

Of the 553 individuals who were emailed the survey, 22.42% (n=124) completed it. We removed 10 participants for analyzing the data as they had answered only demographics question.

Demographic details of these participants are provided in Table 6. forty-seven (41.1%) participants were female and 66 (57.9%) were male. one participant did not respond to the question concerning gender (0.9%). Most of the participants were between 35 and 44 years of age, 91.4% (n=70 of 114) had a PhD degree, Of the 114 participants who answered the question about their research role, slightly more than a quarter of them 36.8% (n=42) were staff (manager, associate, assistant). Approximately one in three 32.5% (n=37 of 114) were trainees (e.g., MSc., PhD students, postdoctoral fellow).

**Table 6.** Demographic data

| Demographic data |  | N | % |
| --- | --- | --- | --- |
| **Gender**<br>Answered(n=114)<br>Skipped(n=0) | Female | 47 | 41.2% |
|  | Male | 66 | 57.9% |
|  | Prefer not to say | 1 | 0.9% |
| **Age**<br>Answered(n=114)<br>Skipped(n=0) | 18-24 | 10 | 8.8% |
|  | 25-34 | 31 | 27.2% |
|  | 35-44 | 34 | 29.8% |
|  | 45-54 | 17 | 14.9% |
|  | 55-64 | 19 | 16.7% |
|  | 65 or older | 3 | 2.6% |
| Degrees<br>Answered(n=114)<br>Skipped(n=0) | Bachelor's degree | 14 | 12.3% |
|  | Master's degree | 21 | 18.4% |
|  | MD | 6 | 5.3% |
|  | PhD | 70 | 61.4% |
|  | Prefer not to say | 1 | 0.9% |
|  | Other (please specify) | 2 | 1.8% |
| Status at The Neuro<br>Answered(n=114)<br>Skipped(n=0) | Trainee (e.g., Msc, PhD student, Postdoctoral fellow) | 37 | 32.5% |
|  | Staff (manager, associate, assistant) | 42 | 36.8% |
|  | Principal Investigator | 33 | 28.9% |
|  | Prefer not to say | 2 | 1.8% |

#### Data sharing practices and Training in data sharing

We asked participants two questions about their data sharing practices in the past 12 months (see table 7). Over half of those who responded 61.4% (n=70 of 114) reported that they published a first or last authored paper in the last 12 months. Of these, 70.0% (n=49) indicated that they openly shared research data related to one of their publications.

**Table 7.**
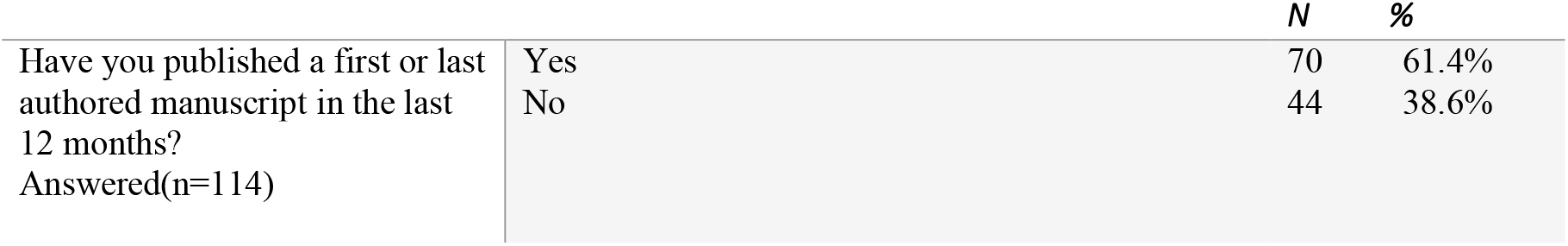
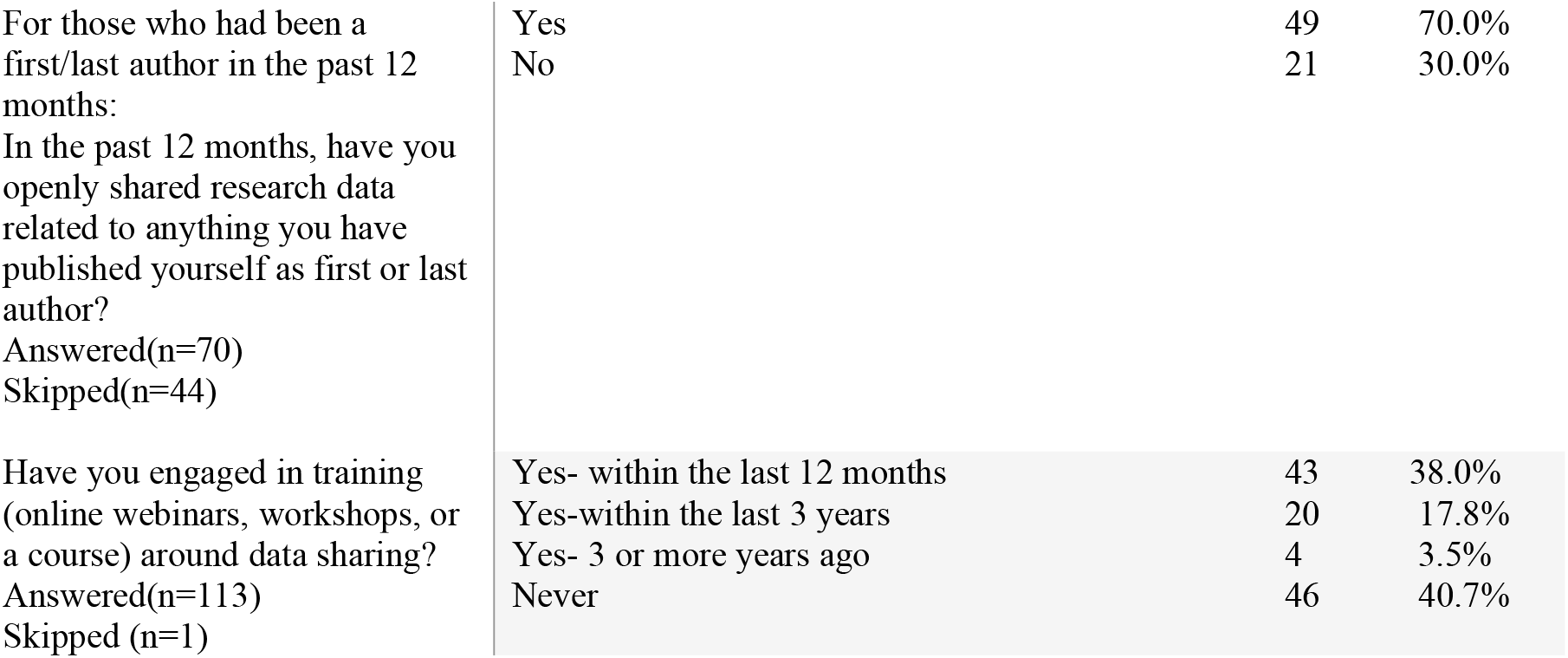
Data sharing experience and training

We asked one question about past and future engagement in training about data sharing. The results are presented in Table 7. Approximately 40% of the respondents indicated that they had never engaged in training related to data sharing (e.g., online webinars, workshops, or a course) (n=49 of 113). Approximately a third of the respondents (38%; n=43) reported having engaged in some form of training (e.g., online webinars, workshops, or a course) around data sharing within the last 12 months. Also, twenty (17.8%) respondents reported having engaged in training (online webinars, workshops, or a course) around data sharing within the last three years and few respondents 3.5% (n=4) indicated they engaged in training around data sharing more than three years ago.

Participants indicated a preference for an online training video that they could return to and a series of several modules, each lasting about 10 minutes and a live webinar were less preferred by participants. These results are reported in Table 8

**Table 8:**
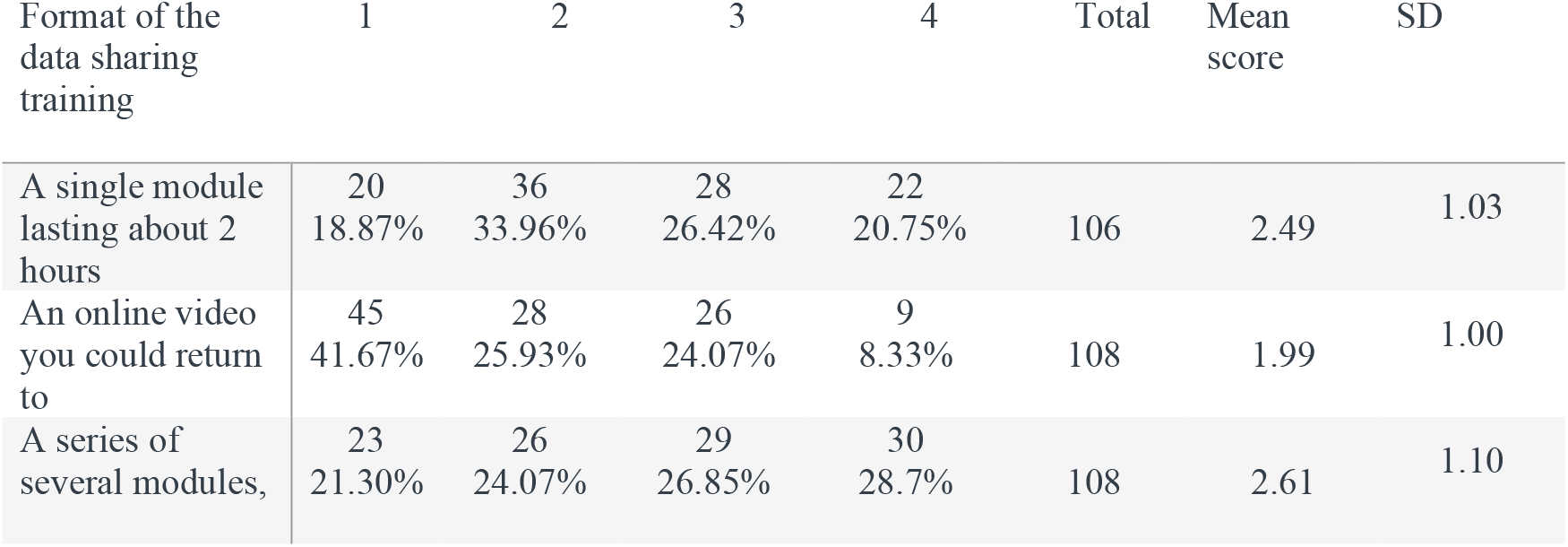
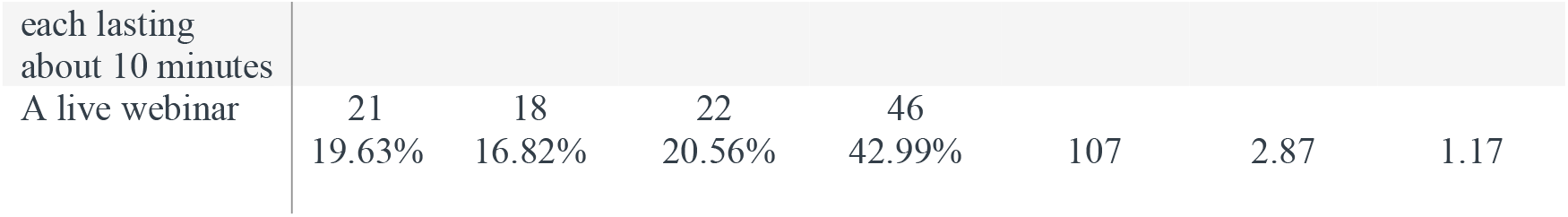
Preferences for data sharing training format

Participants showed a preference for an online handbook walking through the practical steps of data sharing. Next, they indicated they would value having access to a data sharing expert (hired by the Neuro) to directly consult with when questions arise about data sharing. The order of the preference for other resources was: an online interactive learning module, an online video walking through the practical steps of data sharing (why, where, how), a central data sharing expert that facilitates data sharing for projects working directly with the project team, a collection of best practice case study examples. See table 9

**Table 9.** What types of data sharing resources would be most helpful for you? (scale=6)

| What types of data sharing resources would be most helpful for you? | 1 | 2 | 3 | 4 | 5 | 6 | Total | Mean Score | SD |
| --- | --- | --- | --- | --- | --- | --- | --- | --- | --- |
| An online interactive learning module | 22<br>20.95% | 17<br>16.19% | 16<br>15.24% | 15<br>14.292% | 12<br>11.43% | 23<br>21.90% | 105 | 3.45 | 1.84 |
| An online handbook walking through the practical steps of data sharing (why, where, how) | 27<br>25.47% | 20<br>18.87% | 22<br>20.75% | 17<br>16.04% | 14<br>13.21% | 6<br>5.66% | 106 | 2.90 | 1.55 |
| An online video walking through the practical steps of data sharing (why, where, how) | 11<br>10.38% | 19<br>17.92% | 25<br>23.58% | 23<br>21.70% | 14<br>13.23% | 14<br>13.21% | 106 | 3.49 | 1.52 |
| Access to a data sharing expert (hired by the Neuro) to directly consult with when questions arise about data sharing | 22<br>20.75% | 24<br>22.64% | 13<br>12.26% | 24<br>22.64% | 13<br>12.26% | 10<br>9.43% | 106 | 3.11 | 1.62 |
| A collection of best practice case study examples | 7<br>6.60% | 6<br>5.66% | 23<br>21.70% | 16<br>15.09% | 29<br>27.36% | 25<br>23.58% | 106 | 4.22 | 1.49 |
| A central data sharing expert that facilitates data sharing for projects working directly with the project team | 18<br>16.82% | 21<br>19.63% | 8<br>7.48% | 10<br>9.35% | 23<br>21.50% | 27<br>25.23% | 107 | 3.75 | 1.89 |

#### Perceptions about data sharing

Participants were most familiar with patient privacy considerations when sharing data (Mean=3.14, SD=1.091), the ethical considerations when sharing their data (Mean=3.13, SD=1.001) and practical steps involved to share their data (Mean= 2.91, SD=1.085). Respondents were less familiar with concepts including First Nations Principles of Ownership, Control, Access, and Possession (OCAP) (7) (Mean=1.45 =, SD=1.710) and new metrics to measure data sharing contributions (Mean=1.86, SD=0.872). See Table 10 for full results.

**Table 10.** Familiarity with data sharing concepts (Not at all familiar=1, Not so familiar=2, Somewhat familiar=3, Very familiar=4, Extremely familiar=5)

| Data sharing concepts | Not at all familiar | Not so familiar | Somewhat familiar | Very familiar | Extremely familiar | Total | Weighted Average | SD |
| --- | --- | --- | --- | --- | --- | --- | --- | --- |
| Federal mandates related to sharing your data | 35<br>31.53% | 27<br>24.32% | 28<br>25.23% | 19<br>17.12% | 2<br>1.80% | 111 | 2.33 | 1.147 |
| Legal barriers related to sharing your data | 18<br>16.22% | 35<br>31.53% | 42<br>37.84% | 14<br>12.61% | 2<br>1.80% | 111 | 2.52 | 0.971 |
| The new metrics to measure data sharing contributions | 45<br>40.54% | 42<br>37.84% | 20<br>18.02% | 3<br>2.70% | 1<br>0.90% | 111 | 1.86 | 0.872 |
| Ethical considerations when sharing your data | 10<br>9.01% | 13<br>11.71% | 47<br>42.34% | 35<br>31.53% | 6<br>5.41% | 111 | 3.13 | 1.001 |
| Copyright considerations when sharing your data | 16<br>14.41% | 31<br>27.93% | 41<br>36.94% | 17<br>15.32% | 6<br>5.41% | 111 | 2.69 | 1.068 |
| Patient privacy considerations when sharing data | 10<br>9.01% | 18<br>16.22% | 41<br>36.94% | 31<br>27.93% | 11<br>9.91% | 111 | 3.14 | 1.091 |
| The FAIR Principles for data sharing | 39<br>35.14% | 27<br>24.32% | 23<br>20.72% | 18<br>16.22% | 4<br>3.60% | 111 | 2.29 | 1.209 |
| The First Nations Principles of OCAP | 72<br>64.86% | 30<br>27.03% | 8<br>7.21% | 0<br>0.00% | 1<br>0.90% | 111 | 1.45 | 1.710 |
| Practical steps involved to share your data | 16<br>14.68% | 14<br>12.84% | 50<br>45.87% | 22<br>20.18% | 7<br>6.42% | 109 | 2.91 | 1.085 |

Most of the respondents expressed their opinion that data sharing helps to stimulate new hypotheses from the study (Mean=4.38, SD=0.671), they want to help others to use their study results (Mean=4.34, SD=0.691), data sharing helps the advancement of their research by allowing additional investigators to access the data for future research (Mean=4.33, SD=0.730), that they want to help others to reproduce their study (Mean=4.31, SD=0.767), that they want to help others to transparently assess their study (Mean=4.28, SD=0.720), and that they are optimistic that efforts to adhere to data sharing best practices will help support greater reproducibility and transparency of research (Mean=4.27, SD=0.862). Most of the respondents think that the benefit of data sharing is delayed and uncertain (Mean=2.51, SD=0.989). Also, most respondents disagreed that there is sufficient financial support to help them to adhere to data sharing best practices in the coming year (Mean=2.57, SD=1.016), that they feel stressed out when they think about how to adhere to best practices regarding data sharing for their studies in the coming year. (Mean=2.72, SD=1.014). For full results, see Appendix 3

#### Thematic analysis

For the item “What incentives do you think the Montreal Neurological Institute-Hospital could introduce to recognize data sharing?” we classified text-based responded into six themes. The themes were: 1) financial support, 2) recognize and incentivize data sharing, 3) provide infrastructure to support data sharing, 4) enforcement of specific and clear data sharing standards, 5) change research and recognition priorities, and 6) provide educational support for data sharing (see table 11).

**Table 11.**
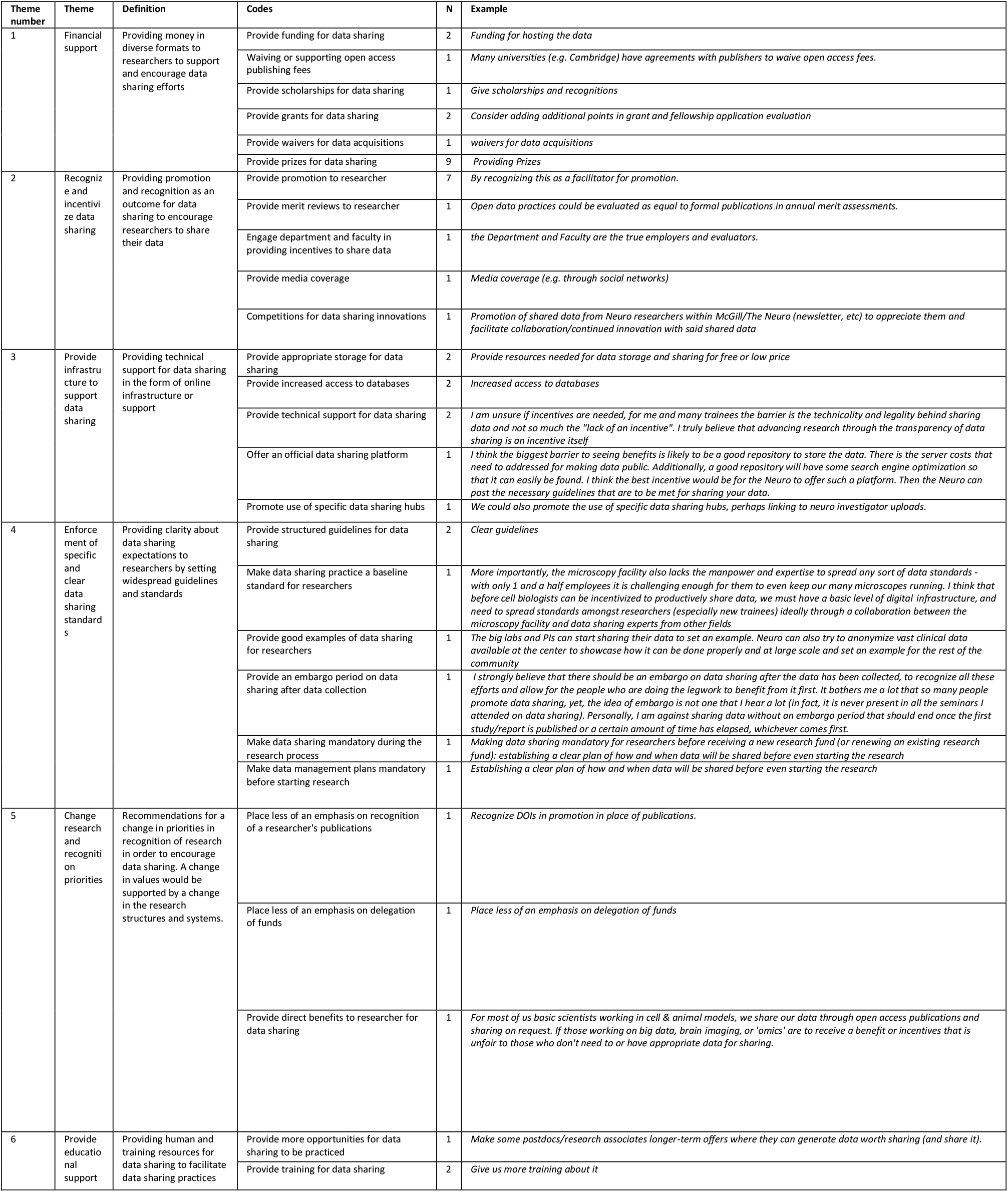
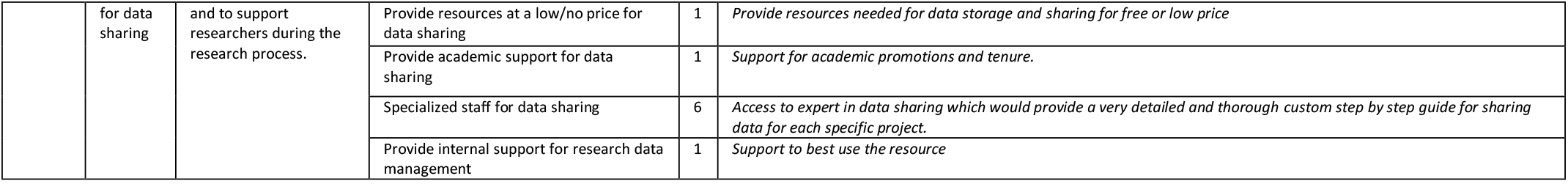
Incentives to recognize data sharing

For the question “Is there anything else you want to share about data sharing?” we classified responses into eight themes. The themes were: 1) lack of resources for data sharing, 2) barriers to data sharing, 3) recommendations for standards surrounding data sharing, 4) relevancy of data sharing, 5) provide training for data sharing, 6) privacy considerations when sharing data, 7) technical requirements for data sharing, 8) data sharing requires more resources (see table 12).

**Table 12.** Comments about data sharing

| N | Theme | Definition | Codes | N | Example |
| --- | --- | --- | --- | --- | --- |
| 1 | Lack of resources for data sharing | Difficulty for data sharing to be put in practice because researchers lack time and resources, which would be put in better use for other aspects of their research | Lack of time for data sharing | 2 | *But it's hard to find the time to do data sharing* |
|  |  |  | Lack of resources for data sharing | 2 | *Weird for an Open Science institution: there is no shared catalog of resources (mouse lines, cell lines, plasmids, equipment, etc..) available at the Neuro.* |
| 2 | Barriers to data sharing | Barriers existing to prevent and discourage researchers from data sharing | Publishing barriers to sharing data | 1 | *The other critical issue that prevents data sharing for the purpose of reproducibility and transparency is the lack of journals that accept purely reproductive or replication research (i.e. studies trying to replicate previous fundings) or journals that will publish negative findings. These are majore barriers to promoting good science and encouraging people to share their data.* |
|  |  |  | Technical barriers prevent data sharing | 3 | *The "technical" barriers I face aren't (e.g.,) knowing how to upload data; it's that hosting our enormous raw data files is expensive and not supported by OSF: 5 Gb is literally 5 minutes of our experiments. A "career" issue I face is that we often collect datasets that let us answer several questions simultaneously and sharing that seems like a recipe to be scooped. I also don't think the incentives are really aligned: an additional paper is far, far more valuable to ECRs than a citation.* |
|  |  |  | Research Ethics Board constrains data sharing | 3 | *The main barrier to data sharing in my work is much REB constraints (on whether data can be shared or not, and which repositories are acceptable for hosting data), and that some of my work is in patients for which data sharing is much more sensitive and hard to get REB-approved.* |
|  |  |  | Ownership/misuse of data concerns when sharing data | 1 | *I'm a research assistant, so I have not my own "study data" to share. I agree that data sharing will help accelerate scientific progress but privacy and personal data sharing must be regulated.* |
|  |  |  | Involvement of the legal system in scientific research is a burden | 1 | *I think the involvement of the legal system and especially lawyers into the mix of scientific inquiry have been a burden and will continue to be. They are a millstone around the neck of scientific progress.* |
|  |  |  | Lack of incentive to share data | 3 | *We receive a lot of training opportunities on data sharing. But it's hard to find the time to do data sharing. And there is neither necessity to do it, nor any major incentive.* |
| 3 | Recommendations for standards surrounding data sharing | These items should be considered when widely enforcing and encouraging data sharing in research | Data sharing standards must accommodate diversity in research | 2 | *I don't do brain imaging for which there are a lot of standards. Transcriptomic data is easy to share because there are repositories for this and so this data is easy for me to share because there are clear instructions. Other data types, microscopy imaging, flow cytometry, MEA do not have clear standards or repositories (that I am aware of). I would* *like to share data but I don't know how. I share my code on github, but I don't know if I'm doing thing correctly.* |
|  |  |  | Data sharing requires innovation | 1 | *we need to innovate, not just be followers.* |
|  |  |  | Data sharing standards must accommodate diversity in research | 1 | *Guidelines for adequate data sharing practices should be introduced to all Neuro labs. There are waste differences in the type of research (thus data produced) conducted at the Neuro, that should be openly shared however labs need to know the recommended best practices: which omics repositories to use, Western blot repositories, when to publish on pre-print servers, etc.* |
|  |  |  | Provide guidelines for data sharing | 1 | *Doing it correctly is not clear and having explicit online guidelines devoid of jargon and preassumptions would be great. I find that I cannot even get started often because the resources assume greater expertise in database management than I have.* |
| 4 | Relevancy of data sharing | Data sharing can feel more or less important depending on a researcher's specific field or who may benefit from data sharing | Data sharing is not relevant to my research | 3 | *Because my lab doesn't study people, my answers don't reflect any concerns about privacy* |
|  |  |  | Data sharing should be more accessible to researchers and the public | 1 | *However the general findings and how they are applicable for the public at large is important and I feel is lost in the process of publication.* |
| 5 | Provide training for data sharing | Providing training and education about data sharing practices can encourage the practice of data sharing | Provide training for data sharing | 3 | *I'm a new master's student and I feel like I haven't been exposed to any data sharing information sessions. I'm at the point where I'm only beginning to generate data so sharing it isn't something that I've even thought of. I have almost zero knowledge on how to go about it. It could be nice to have some training on for new students so we can have these ideas in mind from the start of our careers in science.* |
|  |  |  | Researchers need clarity on when to share data | 1 | *I think the exact moment when sharing has to be carefully considered.* |
| 6 | Privacy considerations when sharing data | There exist concerns when sharing data in specific research (e.g., patient data in clinical trials) | Privacy concerns when sharing data | 2 | *But privacy and personal data sharing must be regulated.* |
|  |  |  | Data sharing privacy concerns are not relevant to all research | 1 | *privacy and personal data sharing must be regulated.* |
| 7 | Technical requirements for data sharing<br>Data sharing requires more resources | Providing support for the technical requirements of data sharing is necessary for data sharing practices to be facilitated | Provide the required infrastructure for data sharing | 2 | *We need tools and infrastructure that actually work and are inclusive of all research realities.* |
|  |  |  | Provide repositories for data sharing | 1 | *Guidelines for adequate data sharing practices should be introduced to all Neuro labs. There are waste differences in the type of research (thus data produced) conducted at the Neuro, that should be openly shared however labs need to know the recommended best practices: which omics repositories to use, Western blot repositories, when to publish on pre-print servers, etc.* |
|  |  |  | Provide technical support for data sharing | 3 | *It's more complicated than you think. The Neuro needs to put more support into the technical side of open data-sharing. Of course, the 'marketing' and administration of open neuroscience at the Neurologist is important but without a solid technical basis, the entire enterprise will fail.* |
| 8 | Data sharing requires more resources | Requirements for human or financial resources in order for data sharing to be practiced | Data sharing requires more funding | 2 | *Open Access is great and needs to be championed, however it is unreasonable to expect that every lab a) can do it, or b) should, due to tight funding and unethically competitive practices of other labs in the field* |
|  |  |  | Provide internal support for data sharing | 1 | *The Neuro needs to put more support into the technical side of open data-sharing* |

## Discussion

The aim of the first study was to audit all publications produced by Neuro researchers in 2019. We found that 66.5% (n=208) of publications had a data sharing statements and 61% (n=31) of publications specified to contact the author to gain access to the data. Sharing by request creates burden and barriers for other researchers to access and use data and should only be implemented in instances where open data is not possible. Most of the publications indicated that there is no registration statements and protocol availability. We note that the authors are more eager to share their publications material than analysis scripts. Most publications did not report using a reporting guideline to maximize the transparency and completed of their research.

In the second study, to identify barriers and facilitators to data sharing, we found that there was a preference that training in the form of an online video they could return to, a series of several modules, each lasting about 10 minutes and a single module lasting about 2 hours be developed. A key finding was that more than a third of the respondents (40.7%), had not engaged in training around data sharing. We identified learning gaps in some key areas (e.g., OCAP Principles, metrics to measure data sharing contributions). Respondents noted that barriers they faced when sharing data included: financial support, training, and technical support.

We recommend an educational and training intervention devoted to data sharing practices to further normalize and support this practice, including training on sharing statistical analysis plans and sharing of other materials related to data sharing. This training could then be followed by additional sessions on further open science practices. Further research is needed to examine the universities’ data sharing practices to track needs and preferences over time and investigate to provide clear data sharing standards for the staff, researchers, and managers.

Also, it is important to repeat the audit over time to track changes in the Neuro publications and researcher perceptions and barriers and facilitators. Hence, it is important to repeat the audit biennially with the same questions in survey. It is also should be mentioned that in addition to the same questions, additional questions should be added to gain a better understanding about certain data sharing practices in universities and scientific institutes.

### Strengths and limitations

The results of the two studies will provide the Neuro with a better understanding of open access statues and barriers and facilitators to data management and sharing and will identify educational needs related to data sharing that can be reduce barriers to data sharing. We hope this report also provides other organizations wanting to engage their communities about data sharing with a valid approach to the topic and some comparative data. Further, the audit of publications can be used to benchmark for improvements over time and to monitor change. Also, Decision-makers in government and universities will be able to structure future open access by providing training for their researchers. Also, this study can serve as a baseline to benchmark for improvements in data sharing and other open science practices and to measure progress over time.

We acknowledge certain limitations. First, as less than a quarter of the Montreal Neurological Institute-Hospital’s staff completed this survey, the results of the study cannot be generalized. Also, it is hard to measure changes in the community unless two or more surveys are done in different time periods. Hence, we recommend annual surveys of the Neuro community to track changes in needs and preferences over time.

## Data Availability

All study data and materials have been made publicly available: https://osf.io/swyvc/.

## Ethics Approval Statement

The study received ethical approval from the Ottawa Health Science Research Network Research Ethics Board (OHSN-REB #20210514-01H).

## Registration

This study was registered a priori (i.e., before any data collection) using the Open Science Framework (https://osf.io/3tafc).

## Data availability statement

It has been deposited in a preprint repository (medRxiv). All study data and materials have been made publicly available: https://osf.io/swyvc/.

## Funding statements

This work was supported by the Tanenbaum Open Science Institute (TOSI). There is no grant number associated with this award.

## Competing interests

The authors received funds from Tanenbaum Open Science Institute to complete the project.

## Conflicts of interest

The authors declared no conflicts of interest.

## Reporting guidelines

We referred to the STROBE and CHERRIES reporting guideline.

## Author statement

**Conceptualization:** S. Ebrahimzadeh, K.D. Cobey, D. Moher; **Methodology:** S. Ebrahimzadeh, K.D. Cobey, D. Moher; **Investigation:** S. Ebrahimzadeh, K.D. Cobey, D. Moher; **Writing the first draft**: S. Ebrahimzadeh; **Writing, Review and Editing:** S. Ebrahimzadeh, K.D. Cobey, J. Presseau, M. Alayche and J.V. Willis, D. Moher; **Funding Acquisition:** D. Moher; **Supervision:** K.D. Cobey and D. Moher

## Acknowledgements

We thank Alex Amar-Zifkin for helping us to design the search strategy, Dylan Roskams-Edris for providing information on the Montreal Neurological Institute-Hospital staff lists and for facilitating the circulation of the survey among the Montreal Neurological Institute-Hospital researchers and the open science grassroots committee of the Montreal Neurological Institute-Hospital for providing feedback on our extraction form and the survey.

## Appendices

### Appendix 1. Search strategy

**Web of Science advanced search template:**

(AU=LASTNAME, FIRSTINITIAL*) AND (PY=2019)

To narrow by contents of address field, add AND (AD=(McGill*) OR (AD=Montreal))

Samples:

(AU=Armstrong, G*) AND (PY=2019)

= 120 results – too many for one person in one year; validate by skimming the results and seeing if any are not our person (ie Gavin, Gregory, when The Neuro’s is ‘Gary’). Apply the address filter.

(AU=Armstrong, G*) AND (PY=2019) AND (AD=(McGill*) OR (AD=Montreal))

= 3 results – makes more sense.

**For researchers with multiple names:**

variations does not affect retrieval:

**For example**,

https://www.mcgill.ca/neuro/etienne-de-villers-sidani-md

in the case of Dr. de Villers-Sidani, variations does not affect retrieval:

((AU=De Villers-Sidani, E*) OR (AU=Sidani, E*) OR (AU=De Villers, E*)) AND (PY=2019)

retrieved the same publications as

(AU=De Villers-Sidani, E*) AND (PY=2019)

Conclude the searching by looking for The Neuro as an institution, in case there’s an author or group author whose name isn’t on the list we have.

This query could be:

((OO=montreal* neurol* hosp*) OR (OO=montreal* neurol* inst*)) AND PY=2019 The OO field is ‘organization’, and it looks to be sourced from Address as well.

**MedRxiv and BioRxiv search template:**

MedRxiv and bioRxiv can be searched simultaneously from https://www.medrxiv.org/search

There is not a way to limit or filter by address.

Use year and lastname, firstinitial-asterisk and manually screening down non-Neuro authors.

**Sample query:**

Rouleau G* retrieved ‘Guy Rouleau’ and ‘Guy A. Rouleau’(fig 1).

**Fig. 1.**
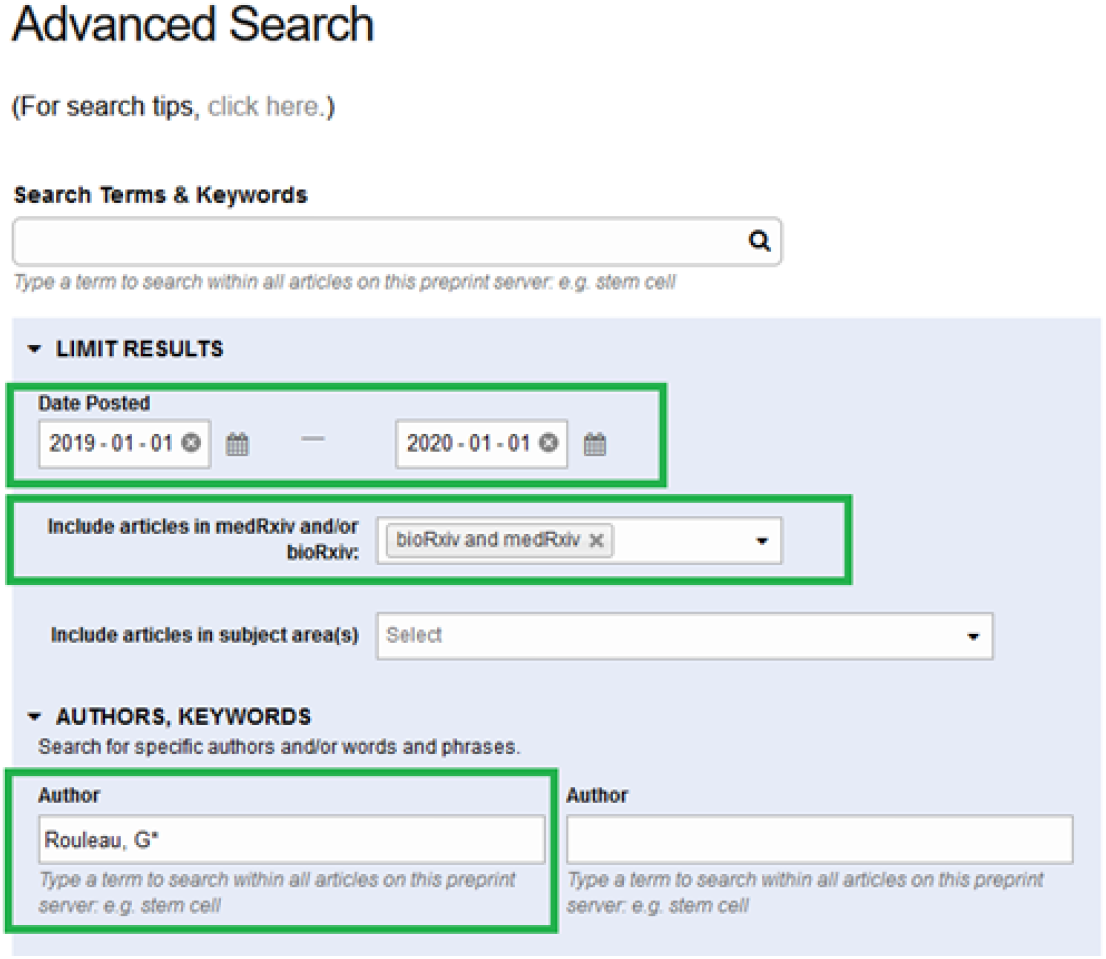
MedRxiv and BioRxiv advanced search (from https://www.medrxiv.org) From https://www.medrxiv.org/content/search-tips:

#### Author Search

1. If a special (i.e., accented) character is included, names with and without the special character will be found (e.g., searching for author Pérez or author Perez will return the same result).
2. Search is performed across both first name and surname.

### Appendix 2. Survey of The Neuro’s Researchers

**Demographic information**

1. What is your current gender identity:

Female

Male

Trans male / Trans man

Trans female / Trans woman

Genderqueer / Gender non-confirming

Other, please specify

Prefer not to say

2. What age group do you fall under?

18-24

25-34

35-44

45-54

55-64

65 or older

Prefer not to say

3. Please select which degrees you have:

Bachelor’s degree

Master’s degree

PhD

MD

Other, please specify

Prefer not to say

4. Which category best describes your status at The Neuro:

Trainee (e.g., Msc, PhD student)

Postdoctoral fellow

Research manager

Research associate

Research assistant

Clinician Scientist

Principal Investigator

Scientist/Independent Researcher

Prefer not to say

**Non-scale items:**

**It should be noted that in this survey data sharing refers to the practice of making data and/or analytical code used to manipulate the underlying data freely available to others to use and build upon**.

5. Have you published a first or last authored manuscript in the last 12 months?

Yes

No

6. In the past 12 months, have you openly shared research data related to anything you have published yourself as first or last author? This means the data was publicly available and others did not need to interact with you to access the data.

Yes

No

7. Have you engaged in training (online webinars, workshops, or a course) around data sharing?

Never

Yes- within that last 12 months

Yes- within the last 3 years

Yes- 3 or more years ago

8. If yes, was training from the Neuro or elsewhere?
9. If you were to engage in training about data sharing what format would you prefer (Rank order)

A single module lasting about 2 hours in

A series of several modules, each lasting about 10 minutes

A live webinar

An online video you could return to

10. What types of data sharing resources would be most helpful for you? Rank order.

An online interactive learning module

An online handbook walking through practical steps of data sharing (why, where, how)

An online video walking through the practical steps of data sharing (why, where, how)

Access to a data sharing expert (hired by the Neuro) to directly consult with when questions arise about data sharing

A collection of best practice case study examples

A central data sharing expert that facilitates data sharing for projects working directly with the project team

The following questions all refer to you personally as a biomedical researcher. Please keep this in mind when responding to the items below. There are no right or wrong answers; we are simply interested in your views

| Domain | Question |
| --- | --- |
| D1. Knowledge | **How familiar are you with each of the concepts listed below?**<br>Federal mandates related to sharing your data<br>(Scale: Not at all familiar, Slightly familiar, Moderately familiar, Very familiar, Extremely familiar)<br>Legal barriers related to sharing your data<br>(Scale: Not at all familiar, Slightly familiar, Moderately familiar, Very familiar, Extremely familiar)<br>Practical steps involved to share your data<br>(Scale: Not at all familiar, Slightly familiar, Moderately familiar, Very familiar, Extremely familiar)<br>Ethical considerations when sharing your data<br>(Scale: Not at all familiar, Slightly familiar, Moderately familiar, Very familiar, Extremely familiar)<br>Copyright considerations when sharing your data<br>(Scale: Not at all familiar, Slightly familiar, Moderately familiar, Very familiar, Extremely familiar)<br>Patient privacy considerations when sharing data<br>(Scale: Not at all familiar, Slightly familiar, Moderately familiar, Very familiar, Extremely familiar)<br>The FAIR Principles for data sharing<br>(Scale: Not at all familiar, Slightly familiar, Moderately familiar, Very familiar, Extremely familiar)<br>The First Nations Principles of OCAP<br>(Scale: Not at all familiar, Slightly familiar, Moderately familiar, Very familiar, Extremely familiar)<br>The new metrics to measure data sharing contributions<br>(Scale: Not at all familiar, Slightly familiar, Moderately familiar, Very familiar, Extremely familiar) |
| D2. Skills | I have the skills needed to identify where and how to share my data (e.g., what platforms to use and how to navigate these)<br>(Scale: Strongly disagree – Disagree – Unsure – Agree – Strongly Agree)<br>I have the skills needed to ensure that the data is share is findable (e.g., uses a persistent identifier, is indexed, is searchable)<br>(Scale: Strongly disagree – Disagree – Unsure – Agree – Strongly Agree)<br>I have the skills needed to ensure my data is shared I way that makes it interoperable (e.g., can easily be integrated with other data, can work with a range of data applications and tools)<br>(Scale: Strongly disagree – Disagree – Unsure – Agree – Strongly Agree)<br>I have the skills needed to prepare and format my data for sharing to ensure data is reusable by others (e.g., know how to format data so that it will be meaningful to other, know how to make a codebook, know how to organize data)<br>(Scale: Strongly disagree – Disagree – Unsure – Agree – Strongly Agree)<br>I have received enough training about how to share my study data when publishing.<br>Scale: Strongly disagree – Disagree – Unsure – Agree – Strongly Agree<br>Most of my skills related to sharing research data have been learned on the job.<br>(Scale: Strongly disagree – Disagree – Unsure – Agree – Strongly Agree) |
| D3.Social/professional role and identity | It is my responsibility as a researcher to adhere to best practices concerning data sharing<br>(Scale: Strongly disagree – Disagree – Unsure – Agree – Strongly Agree) |
| D4. Beliefs about capabilities | I do not feel competent navigating all of the steps to share my study data<br>(Scale: Strongly disagree – Disagree – Unsure – Agree – Strongly Agree)<br>I have control over whether the data from the studies I conduct in the future is shared.<br>(Scale: Strongly disagree – Disagree – Unsure – Agree – Strongly Agree)<br>Sharing of the data is technically easy for me.<br>(Scale: Strongly disagree – Disagree – Unsure – Agree – Strongly Agree)<br>I think the potential benefit of sharing data outweigh the risk such as loss of privacy.<br>(Scale: Strongly disagree – Disagree – Unsure – Agree – Strongly Agree) |
| D5. Optimism | I am optimistic that efforts to adhere to data sharing best practices will help to drive innovation and discovery in biomedicine.<br>(Scale: Strongly disagree – Disagree – Unsure – Agree – Strongly Agree)<br>I am optimistic that efforts to adhere to data sharing best practices will help support greater reproducibility and transparency of research.<br>(Scale: Strongly disagree – Disagree – Unsure – Agree – Strongly Agree) |
| D6. Beliefs about consequences | If I commit to sharing the data from my research studies over the next 12 months, it will...<br>(Scale: Strongly disagree – Disagree – Unsure – Agree – Strongly Agree)<br>Help others to transparently assess my study |
|  | (Scale: Strongly disagree – Disagree – Unsure – Agree – Strongly Agree)<br>Help others to reproduce my study<br>(Scale: Strongly disagree – Disagree – Unsure – Agree – Strongly Agree)<br>Help others to use my study results<br>(Scale: Strongly disagree – Disagree – Unsure – Agree – Strongly Agree)<br>Help others to innovate from my study<br>(Scale: Strongly disagree – Disagree – Unsure – Agree – Strongly Agree)<br>Data sharing helps the advancement of my research by allowing additional investigators to access the data for future research<br>(Scale: Strongly disagree – Disagree – Unsure – Agree – Strongly Agree)<br>Sharing data makes my data broadly accessible.<br>(Scale: Strongly disagree – Disagree – Unsure – Agree – Strongly Agree)<br>I think the benefit of data sharing is delayed and uncertain.<br>(Scale: Strongly disagree – Disagree – Unsure – Agree – Strongly Agree) |
| D7. Reinforcement | If I share my study data for research conducted over the next year, I will get recognition from my institution.<br>(Scale: Strongly disagree – Disagree – Unsure – Agree – Strongly Agree)<br>If I share my study data for research conducted over the next year, I will get recognition from my colleagues whose opinions I value.<br>(Scale: Strongly disagree – Disagree – Unsure – Agree – Strongly Agree) |
| D8. Intention | I intend to share all of my relevant study data over the next year.<br>(Scale: Strongly disagree – Disagree – Unsure – Agree – Strongly Agree)<br>I would be willing to engage in a short online training module about data sharing offered by the neuro<br>(Scale: Strongly disagree – Disagree – Unsure – Agree – Strongly Agree) |
| D9. Goals | It is a high priority for me in the next year to ensure I share all of my relevant study data.<br>(Scale: Strongly disagree – Disagree – Unsure – Agree – Strongly Agree)<br>My first priority in the next year is to learn more about data sharing.<br>(Scale: Strongly disagree – Disagree – Unsure – Agree – Strongly Agree)<br>It is a high priority for me to ensure I am up to date on data sharing best practices.<br>(Scale: Strongly disagree – Disagree – Unsure – Agree – Strongly Agree) |
| D10. Memories, attention, and decision processes | I always remember that I need to share the research data.<br>(Scale: Strongly disagree – Disagree – Unsure – Agree – Strongly Agree)<br>It is easy for me to remember the process of data sharing.<br>(Scale: Strongly disagree – Disagree – Unsure – Agree – Strongly Agree) |
| D11. Environmental context and resources | There are enough resources at my institution to support me in effectively sharing my research data.<br>(Scale: Strongly disagree – Disagree – Unsure – Agree – Strongly Agree)<br>There are sufficient human resources (explicate and implicit knowledge) at my institution to support me in effectively sharing my research data.<br>(Scale: Strongly disagree – Disagree – Unsure – Agree – Strongly Agree)<br>There is sufficient financial support (money or its equivalent) (e.g., CIHR, Institution) to help me to adhere to data sharing best practices in the coming year<br>(Scale: Strongly disagree – Disagree – Unsure – Agree – Strongly Agree)<br>There are enough social resources (networks, norms and values) at my institution to support me in effectively sharing my research data.<br>(Scale: Strongly disagree – Disagree – Unsure – Agree – Strongly Agree)<br>There are transparent guidelines about sharing my research data at my institution.<br>(Scale: Strongly disagree – Disagree – Unsure – Agree – Strongly Agree)<br>There is sufficient technical support to share my research data at my institution.<br>(Scale: Strongly disagree – Disagree – Unsure – Agree – Strongly Agree) |
| D12. Social influences | Researchers whose opinion I value would approve of me committing to sharing all of my study data over the next year.<br>(Scale: Strongly disagree – Disagree – Unsure – Agree – Strongly Agree)<br>Respected researchers in my field of study are sharing their research data.<br>(Scale: Strongly disagree – Disagree – Unsure – Agree – Strongly Agree)<br>My colleagues are familiar with data sharing practices and are willing to help me with it<br>(Scale: Strongly disagree – Disagree – Unsure – Agree – Strongly Agree) |
| D13. Emotions | I feel stressed out when I think about how to adhere to best practices regarding data sharing for my studies in the coming year.<br>(Scale: Strongly disagree – Disagree – Unsure – Agree – Strongly Agree) |
|  | I feel stressed because of the lack of guidelines (e.g., I don't know how to do it confidently)<br>(Scale: Strongly disagree – Disagree – Unsure – Agree – Strongly Agree)<br>I feel satisfied when I spend time sharing my research data.<br>(Scale: Strongly disagree – Disagree – Unsure – Agree – Strongly Agree) |
| D14. Behavioural regulation | Data sharing is something I routinely do automatically and will continue to do so over the next year.<br>(Scale: Strongly disagree – Disagree – Unsure – Agree – Strongly Agree)<br>The culture of openness at the Neuro motivates me to share my study data routinely<br>(Scale: Strongly disagree – Disagree – Unsure – Agree – Strongly Agree) |

#### Text based responses

11. What incentives do you think the neuro could introduce to recognize data sharing?
12. Is there anything else you want to share about data sharing?

### Appendix 3. Agreement with each of the following statements

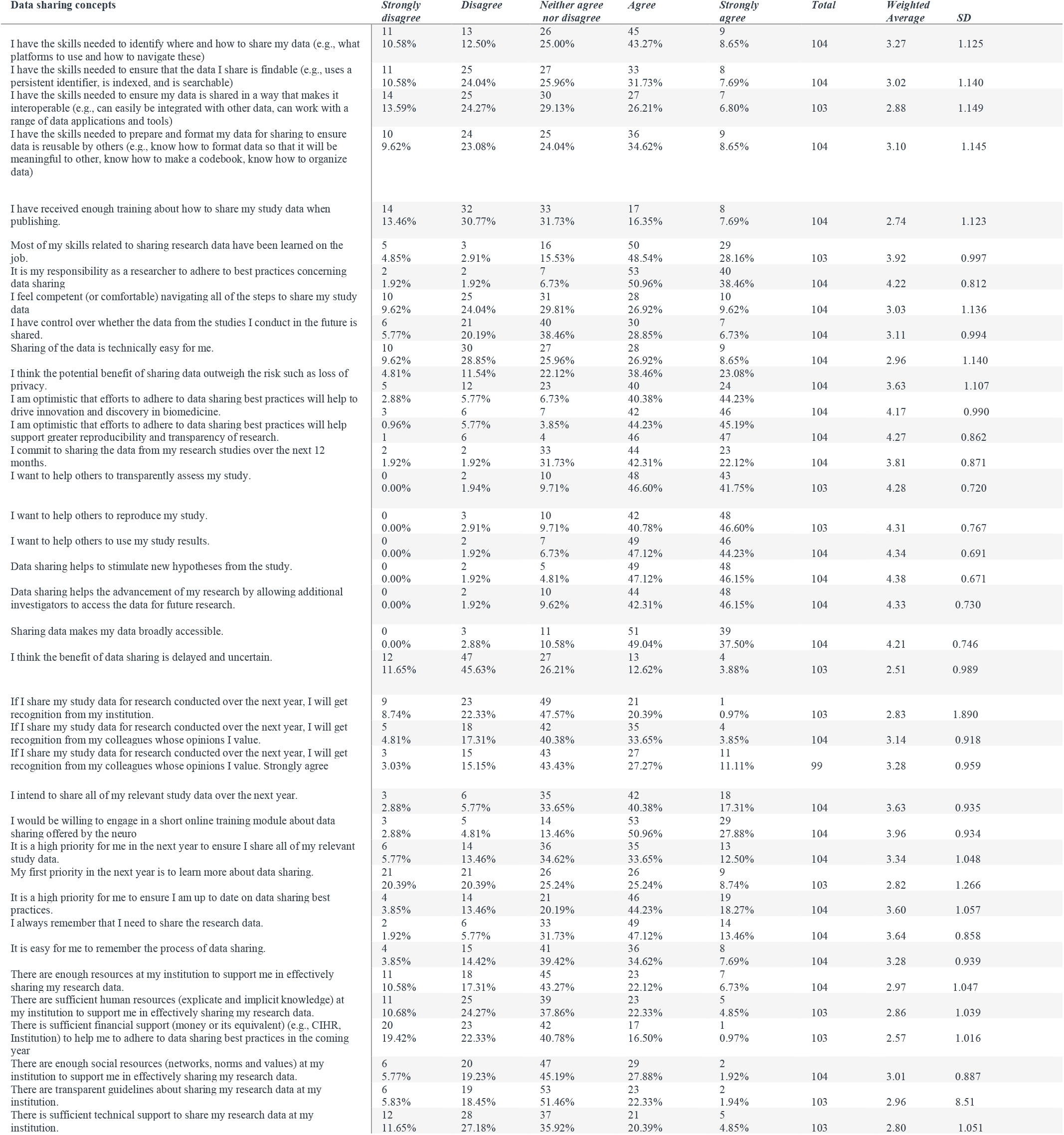
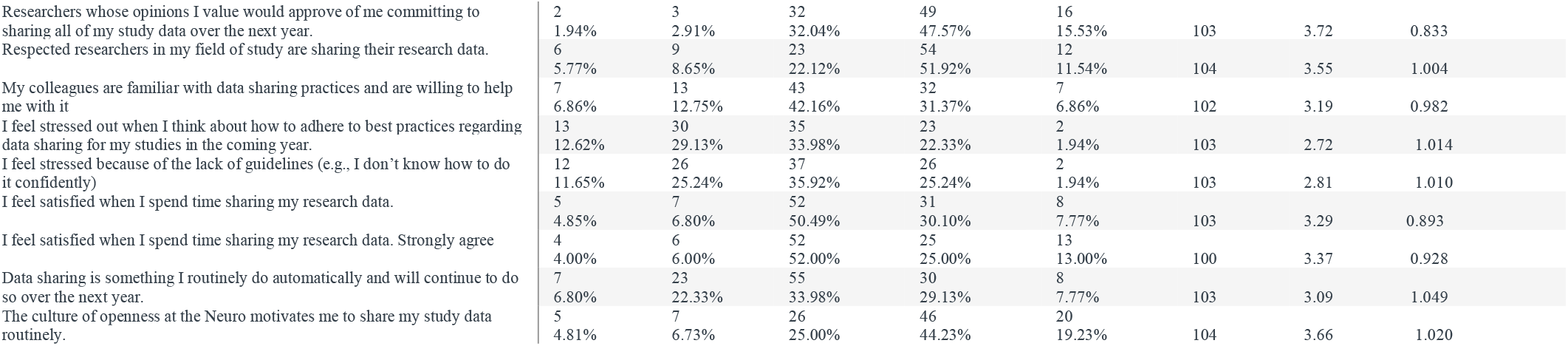

## Notes

### Author Declarations

Ethics committee/REB of Ottawa Health Science Research Network Research Ethics Board gave ethical approval for this work

